# The impact of the COVID-19 pandemic on families in Germany

**DOI:** 10.1101/2020.10.05.20206805

**Authors:** Susanne Brandstetter, Tanja Poulain, Mandy Vogel, Christof Meigen, Michael Melter, Birgit Seelbach-Göbel, Christian Apfelbacher, Wieland Kiess, Michael Kabesch, Antje Körner, the KUNO Kids study group

**Author notes:** These authors contributed equally (Susanne Brandstetter, Tanja Poulain, Michael Kabesch, Antje Körner). **Corresponding author:** Dr. Susanne Brandstetter, University Children’s Hospital Regensburg (KUNO), Hospital St. Hedwig of the Order of St. John, Regensburg, Steinmetzstr., 1-3, 93049 Regensburg, Germany.

## Abstract

**Objective:** To assess the impact of the COVID-19 pandemic on families with young children in two population-based childhood cohorts with a low and moderate COVID-19 prevalence, respectively.

**Methods:** A cross-sectional study using online questionnaires in families from LIFE Child (n=306, Leipzig) and KUNO Kids (n=612, Regensburg) was performed at the end of the German lock-down period. Outcomes were parent-reported impact of the COVID-19 pandemic on family life, concerns and trust in political measures.

**Results:** Most families were concerned about the COVID-19 pandemic and lock-down measures, with major concerns directed towards the economic situation (>70%), the health of close-ones (37%), but less towards their own health (<10%). Many concerns, seeking information and approval of federal measures were more pronounced in the more affected region. Approval of lockdown measures and concerns about economic recession were related to regional differences and not significantly dependent on educational status or being personally affected by the disease.

**Conclusion:** Regional differences in approval of lockdown measures were observed and thus, measures to specifically support families according to the regional impact of the COVID-19 pandemic are needed.

## INTRODUCTION

So far, the COVID-19 pandemic has had very different effects on countries within Europe. France, Spain, Italy and the UK have been most severely affected both in terms of prevalence and mortality rates.^1^ Within Germany, regions in the south and west had much higher rates of COVID-19 cases and deaths than those in the north and east.^2^ However, similar lockdown measures were introduced in all parts of the country. Particularly, closing of daycare nurseries and schools were the first measures taken and the last to be lifted everywhere.^3^

At the beginning of the pandemic, the focus of attention was on elderly people and patients with chronic disease while children were assumed to be not seriously affected by the virus,^3,4^ largely because prevalence rates were thought to be low and severe courses were less frequent in children. Nevertheless, the potential impact on the psychosocial and economic situation of families and the pandemic’s effects on children, such as social deprivation due to the separation from their peers, the absence of education and the potential exposure to violence at home, were only realized later.^5^ Primary data from families affected by the COVID-19 pandemic was however lacking.

In the cities of Leipzig and Regensburg, located in the east (Leipzig) and south (Regensburg) of Germany, two actively recruiting childhood cohorts exist: The LIFE child^6^ and the KUNO Kids health studies.^7^ Therefore, performing a survey during the lockdown phase and asking families about their situation, concerns and constraints during the COVID-19 pandemic were obvious tasks. As children at a young age are particularly vulnerable when social learning develops and peer interaction is key,^8^ the effects of the lockdown may be especially grave.^9^ Thus, we focused on children between 1 and 6 years of age and investigated how families with young children in two regions with considerably different prevalence rates of COVID-19 perceived their situation.

## METHODS

### Design

The study had a cross-sectional design and used data collected in two childhood cohorts in Germany. The cohorts are situated in Leipzig (Saxony) and Regensburg (Bavaria), respectively. Figure 1 provides an overview of the time course of the COVID-19 pandemic and the associated policy measures for both study regions.

**Figure 1:**
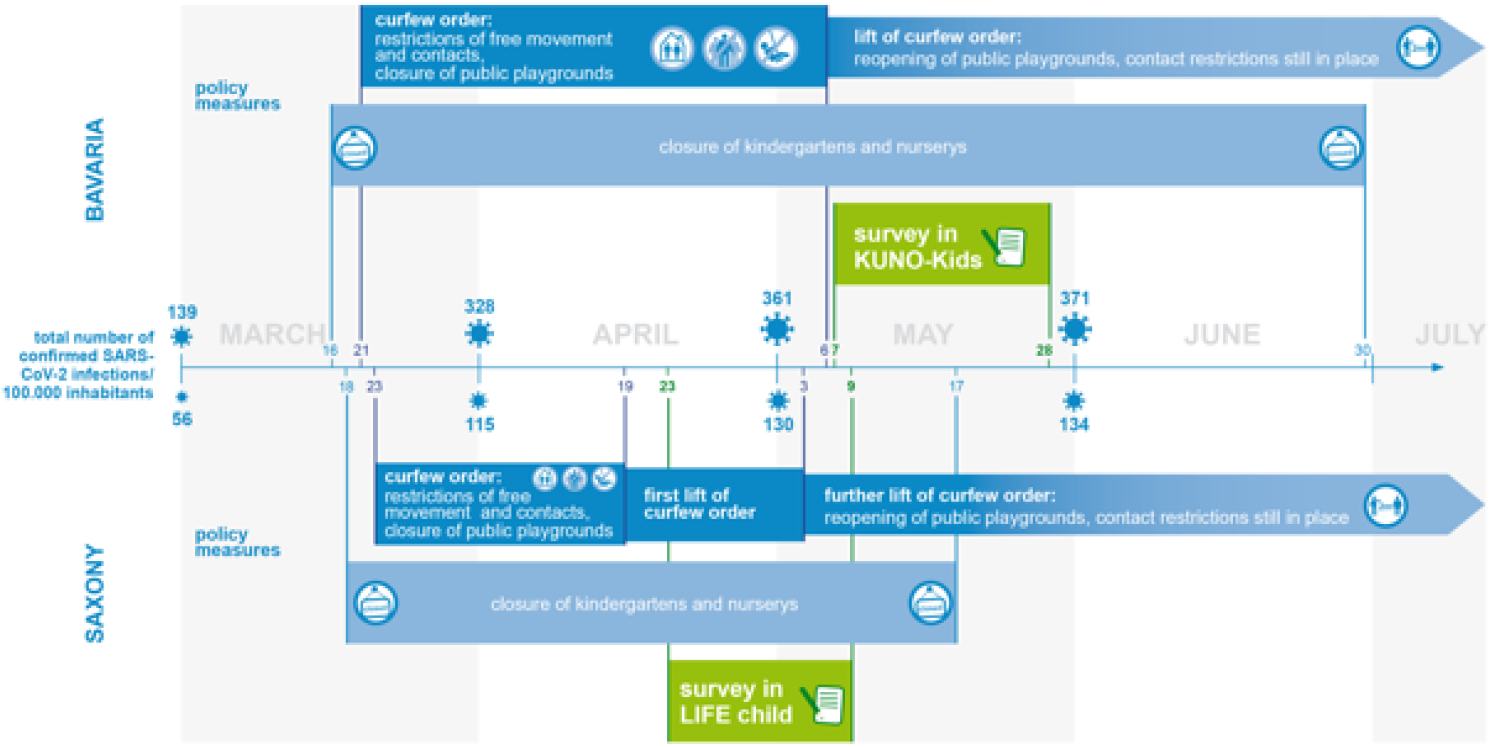
Overview on policy measures and number of SARS-CoV-2 infections in the two states where the study took place: Bavaria (Regensburg) and Saxony (Leipzig)

### Study participants

The LIFE Child study is an ongoing childhood cohort study that started in 2011 and has 4800 participants so far.^6^ The study is conducted in Leipzig, a city with 600.000 inhabitants situated in Eastern Germany. Participants in LIFE Child are recruited from during pregnancy until the age of 16 years and participate in annual examinations. All parents provide written informed consent before participation. With 134 infections/100.000 inhabitants (22.06.2020^2^), Leipzig was mildly affected by the COVID-19 pandemic.

The KUNO-Kids health study is an ongoing, multi-purpose birth cohort which was initiated in 2015 and has 3249 participants at this timepoint.^7^ The study is located in Eastern Bavaria and covers the city of Regensburg (170.000 inhabitants) and the adjacent, mostly rural regions. All mothers giving birth at the clinic St. Hedwig are asked to provide written informed consent for participation in the study. Only one child per family is included in the study. Data are collected after birth of the child and at various follow-ups. Some counties in the area of Regensburg were severely affected by the COVID-19 pandemic with more than 500 SARS-CoV-2 infections/100.000 inhabitants (22.06.2020^10^).

All families with children between the age of 1.5 to 5.9 years, who were currently participating in either LIFE Child study or KUNO-Kids and who had agreed to be contacted for additional studies, were eligible and contacted for the present study. In Leipzig, 721 families were invited to complete the online survey. Of these families, 306 (42.4%) completed the questionnaire between April 23 and Mai 9, 2020. In Regensburg, the invitation was sent to 1296 eligible families. Of these, 612 (50,1%) completed the questionnaire between May 7 and May 28, 2020.

### Questionnaire

The survey comprised questions covering topics relating to the situation of families during the COVID-19 pandemic: SARS-CoV-2 infections in family members or among friends, risk persons among family and friends, current quarantine status and working conditions, coping with the current situation, expectations for the future, various concerns and trust in policy measures.

For both cohorts, data collected in the online survey were complemented by information on age of mother and child, migration background, highest educational level and employment status of both parents, and the child’s visit to a nursery or kindergarten.

### Statistical analyses

Descriptive statistics were reported as frequencies and percentages for dichotomous or ordinal variables, and means (M) and standard deviations (SD) for metric variables, respectively. When describing the survey responses, percentages given in the text refer to the cumulative percentage of the two strongest/worst categories (usually “rather”+”very much”, “somewhat” + “not at all”, respectively). Proportional odds logistic regression analyses were performed to investigate the associations between study region and survey items. In order to analyze associations between predictors and selected outcome variables (regarding concerns and trust) multivariable proportional odds logistic regression analyses were performed. The predictor variables comprised study region (Leipzig vs. Regensburg), sociodemographic variables (education, migration background), the occurrence of SARS-CoV-2 infections in family/friends, risk group members in family/friends and being in quarantine. Predictor variables were simultaneously included in the models. Results are presented by odds ratios (OR) and 95% confidence interval (95% CI). All analyses were performed using R.^11^

## RESULTS

For both cohorts, general characteristics as well as indicators of how participants were affected by the pandemic are shown in Table 1. In 98% of the families, the mother completed the questionnaire. The cohorts did not show relevant differences regarding age of mother or child. The majority of mothers were in permanent employment with children usually attending daycare in both cohorts. Rate of permanent employment of fathers was slightly lower in the Leipzig cohort as was the percentage of migration background. In both cohorts, the majority of parents had a higher educational degree. Most families had close ones belonging to high-risk groups, though only very few families were in actual quarantine. Percentages of close ones with mild/severe COVID-19 were twice as high in the Regensburg cohort corresponding with the marked differences in regional prevalence rates (see also Figure 1). For more than half of the parents, the work situation did not change during the COVID-19 associated lockdown, with about 20% of mothers and 30% of fathers continuing to work from the office.

**Table 1:** Comparison of cohorts and regional differences

|  | LIFE Child |  | KUNO Kids |  |
| --- | --- | --- | --- | --- |
| **n** | 306 |  | 612 |  |
| **Age mother** (years, mean±SD) | 35.4±4.4 |  | 36.3±3.9 |  |
| **Age child** (years, mean±SD) | 3.6±1.3 |  | 3.4±0.9 |  |
| **Child usually in daycare (%)** |  |  |  |  |
| Yes | 61.1 |  | 65.2 |  |
| No | 19.9 |  | 29.6 |  |
| Missing | 19.0 |  | 5.2 |  |
| **Migrational background (%)** |  |  |  |  |
| Yes | 4.2 |  | 11.9 |  |
| No | 71.6 |  | 85.6 |  |
| Missing | 24.2 |  | 2.5 |  |
| **Education parents (%)** |  |  |  |  |
| Lower | 15.0 |  | 21.4 |  |
| Higher | 84.0 |  | 78.1 |  |
| Missing | 1.0 |  | 0.5 |  |
| **Mother in job (%)** |  |  |  |  |
| Yes | 63.7 |  | 66.0 |  |
| No | 34.7 |  | 32.7 |  |
| Missing | 1.6 |  | 1.3 |  |
| **Father in job (%)** |  |  |  |  |
| Yes | 86.3 |  | 94.9 |  |
| No | 6.9 |  | 0.9 |  |
| Missing | 6.8 |  | 4.2 |  |
| **Family members in risk group (%)** |  |  |  |  |
| Yes | 83.7 |  | 85.0 |  |
| No | 16.3 |  | 14.7 |  |
| Missing | 0 |  | 0.3 |  |
| **Current quarantine (%)** |  |  |  |  |
| Yes | 1.6 |  | 0.5 |  |
| No | 98.4 |  | 99.2 |  |
| Missing | 0 |  | 0.3 |  |
| **Confirmed COVID-19 cases in close ones (%)** |  |  |  |  |
| None | 88.6 |  | 75.3 |  |
| Mild | 7.8 |  | 15.2 |  |
| Serious | 3.6 |  | 9.2 |  |
| Missing | 0 |  | 0.3 |  |
| **Impact of lockdown on working situation (%)** |  |  |  |  |
|  | mother | father | mother | father |
| None, same hours | 54.6 | 53.9 | 50.0 | 67.2 |
| None, freelancer | 4.9 | 5.2 | 1.0 | 1.5 |
| Yes, reduced hours | 12.7 | 17.0 | 12.9 | 20.9 |
| Yes, off work | 2.3 | 4.2 | 3.7 | 2.0 |
| Yes, freelancer | 2.9 | 4.9 | 2.3 | 2.9 |
| Lost job | 0.3 | 1.0 | 0.7 | 0.2 |
| Not applicable | 22.0 | 13.1 | 27.3 | 2.9 |
| Missing | 0.3 | 0.7 | 2.1 | 2.4 |

| Home office due to lockdown (%) |  |  |  |  |
| --- | --- | --- | --- | --- |
|  | mother | father | mother | father |
| Yes, completely | 22.2 | 27.4 | 19.6 | 26.8 |
| Yes, partly | 15.0 | 14.0 | 13.6 | 16.7 |
| No | 36.7 | 36.3 | 22.5 | 43.6 |
| No, already home office before | 3.3 | 3.3 | 3.4 | 3.6 |
| Not applicable | 22.5 | 18.3 | 37.1 | 4.9 |
| Missing | 0.3 | 0.7 | 3.8 | 4.4 |

“Child in daycare” refers to the situation before the COVID-19 pandemic; “Migration background” in mother and/or father

The distribution of all the participants’ responses to the survey questions is depicted in Figure 2. Very few families did not care much about the COVID-19 pandemic (12.5%), whereas 29% were “very much” or “extremely” concerned about how things would continue across both cohorts.

**Figure 2:**
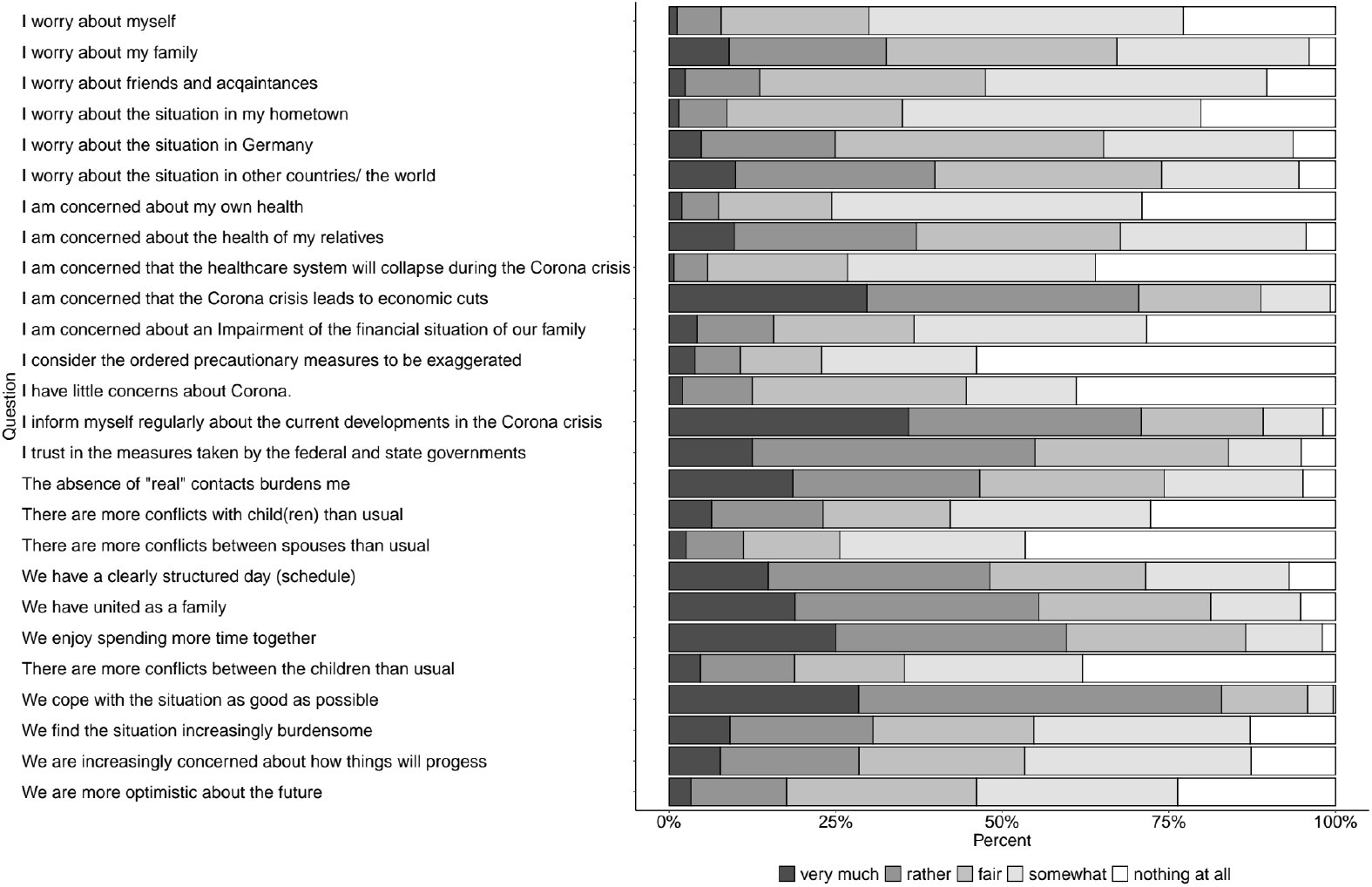
Distribution of participants’ answers to the survey questions (N=918)

When asked what the concerns were directed to, less than 10% of responders were concerned about their very own situation, whereas 40% were concerned about the situation in other countries and the world. Similarly, for health-related concerns, less than 8% worried about their own health, but 37% were concerned about the health of close-ones. The most prevalent concern across all topics was for general economic recession (>70% at least “very much” concerned), though only 15% of families were very much worried that the COVID-19 pandemic affected their own financial situation. Only few families worried about the health care system.

Most responders expected that the situation would normalize within half a year (59.9%), but 37.3% were seriously afraid it would never be the same. More than half of the families trusted very much in the federal measures against COVID-19 pandemic.

Many participants reported an impact of the COVID-19 pandemic on social and family life. About one third of the families (30.5%) were at least very much burdened by the situation, and the lack of social contacts was very/extremely bothersome for almost half of families. On the other hand, the majority coped well with the situation. About half had a clear daily schedule. Families indicated that they were enjoying the additional time they could spend together (60%) and that they had united as a family (56%). Nevertheless, 22.5% of the families reported relevantly more conflicts with children and 19% and 12% reported more conflicts between children or between the spouses, respectively.

The participants’ responses regarding concerns and perceived impact of the COVID-19 pandemic differed between the study regions (Figure 3). Overall concerns were stronger in Regensburg - the region with higher prevalence of SARS-CoV-2 infections. Participants from the Leipzig cohort were less likely to seek information about the pandemic, more likely to not care much about the pandemic and more likely to consider the policy measures as exaggerated.

**Figure 3:**
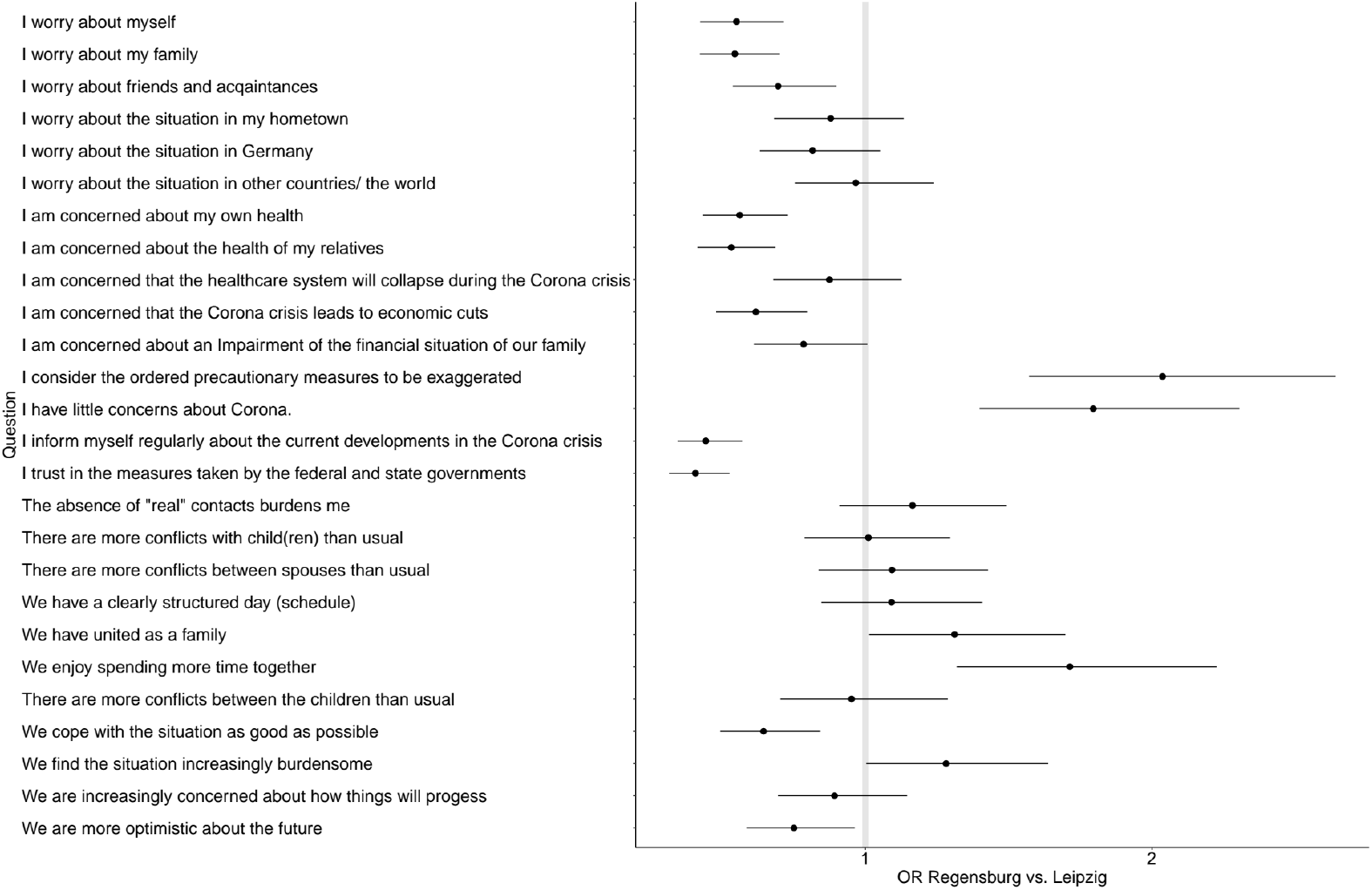
Regional differences in participants’ answers to the survey questions (N=918)

Furthermore, general and health concerns for the family and oneself were almost twice as strong in Regensburg, whereas the concerns for the situation in the hometown/ Germany/ world did not differ between the cohorts. Also, concerns about economic recession, that were very strong in both cities, were even more pronounced in the Regensburg cohort.

Families in Leipzig tended to experience the situation even more burdensome than in Regensburg and stated they coped less well, even though they stated significantly more often that they appreciated more time and cohesion within the families.

Table 2 shows the results of the multivariable analyses. Worries about oneself, about families, about friends and about one’s hometown were significantly stronger if family members or friends belonged to the risk group. Independent of this, families in Leipzig worried less. Worries about the entire world were less intense in participants with lower educational degree and with migration background and stronger if close ones suffered from severe infection. The trust in political measures against the pandemic and concerns about economic recession was significantly lower in Leipzig.

**Table 2.** Effects of different predictors on COVID-19-related worries and concerns estimated by multivariable proportional odds logistic regression.

|  | Worries about oneself |  | Worries about family |  | Worries about friends |  | Worries about hometown |  | Worries about Germany |  | Worries about world |  | Trust in political measures |  | Concerns about economic recession |  |
| --- | --- | --- | --- | --- | --- | --- | --- | --- | --- | --- | --- | --- | --- | --- | --- | --- |
|  | OR (95% CI) | p | OR (95% CI) | p | OR (95% CI) | p | OR (95% CI) | p | OR (95% CI) | p | OR (95% CI) | p | OR (95% CI) | p | OR (95% CI) | p |
| Education (low) | 1.36 (0.99-1.88) | .056 | 1.15 (0.84-1.57) | .379 | 1.24 (0.90-1.70) | .180 | 0.87 (0.63-1.20) | .387 | 0.91 (0.66-1.24) | .538 | **0.52 (0.38-0.72)** | **<.001** | 0.73 (0.52-1.01) | .055 | 1.23 (0.90-1.69) | .197 |
| Migration (yes) | 1.17 (0.76-1.79) | .475 | 1.22 (0.80-1.86) | .344 | 0.98 (0.64-1.52) | .941 | **1.59 (1.02-2.49)** | **.041** | 0.92 (0.60-1.41) | .693 | **0.61 (0.40-0.93)** | **.021** | 0.94 (0.61-1.45) | .770 | 0.87 (0.58-1.32) | .518 |
| Quarantine (yes) | 0.29 (0.76-1.25) | .107 | 1.08 (0.28-3.99) | .910 | 0.52 (0.13-2.05) | .353 | 0.84 (0.19-3.94) | .724 | 0.80 (0.19-3.20) | .751 | 0.77 (0.21-2.89) | .696 | 1.52 (0.26-8.47) | .640 | 0.51 (0.10-2.51) | .410 |
| COVID-19 in close ones (mild) | 1.05 (0.72-1.53) | .791 | 1.08 (0.75-1.57) | .664 | 1.06 (0.73-1.55) | .743 | 1.01 (0.70-1.48) | .944 | 0.83 (0.58-1.20) | .327 | 1.12 (0.78-1.62) | .550 | 1.31 (0.90-1.91) | .0156 | 1.03 (0.71-1.51) | .864 |
| COVID-19 in close ones (serious) | 0.95 (0.58-1.56) | .838 | 0.99 (0.61-1.60) | .977 | 1.50 (0.93-2.43) | .096 | 1.27 (0.79-2.04) | .326 | 1.57 (0.96-2.59) | .073 | **1.83 (1.12-3.01)** | **.017** | 0.89 (0.54-1.46) | .631 | 1.39 (0.85-2.27) | .189 |
| Risk group (yes) | **1.55 (1.08-2.23)** | **.017** | **2.54 (1.78-3.64)** | **<.001** | **1.54 (1.08-2.20)** | **.017** | **1.49 (1.04-2.14)** | **.032** | 1.01 (0.71-1.43) | .961 | 0.79 (0.56-1.11) | .174 | 1.41 (0.99-2.01) | .057 | 1.14 (0.79-1.64) | .486 |
| Region (Leipzig) | **0.54 (0.40-0.72)** | **<.001** | **0.57 (0.43-0.76)** | **<.001** | **0.72 (0.53-0.96)** | **.026** | 0.90 (0.67-1.20) | .473 | 0.78 (0.58-1.04) | .095 | 0.85 (0.64-1.13) | .266 | **0.36 (0.27-0.48)** | **<.001** | **0.63 (0.47-0.84)** | **.002** |
| R<sup>2</sup> | .305 |  | .325 |  | .269 |  | .279 |  | .277 |  | .309 |  | .331 |  | .286 |  |

Multivariable proportional odds logistic regression analyses of COVID-19-related concerns as dependent variable (headings) in = 887 families with data for independent variables: Education: Highest school degree of parents (reference = higher). Migration: At least one parent was not born in Germany (reference = no). Quarantine: Family (member) is currently in quarantine (reference = no). Covid-19 in family/friends (reference = no). Risk group: family members in risk group (reference = no).

## DISCUSSION

During the first stage of the COVID-19 pandemic in Europe, Regensburg experienced infection rates approximately three times as high as those in Leipzig. Measures to contain SARS-CoV-2 infections were quite similar in timing and nature in both regions and affected families with children strongly in their daily life. Most families were concerned about the COVID-19 pandemic and the impact of lock-down measures, with major concerns directed towards economic situation and the lack of social contacts, but less towards their own health. Overall, parents’ concerns, need for information on the pandemic as well as the approval of federal measures were stronger in the more affected region.

This study is based on a survey conducted in two well-characterized cohorts of families with young children. In terms of socio-demographic characteristics the cohorts were largely comparable. By implementing identical questions in both cohorts our study succeeded in (a), achieving a large sample size, which allowed for multivariable analyses, and in (b), comparing regional differences across Bavaria and Saxony. By the timely collection of data attitudes and concerns regarding the COVID-19 pandemic our study explored the situation of families with young children at a crucial point in time. Families had to cope with the restriction measures for many weeks and it was not yet clear when and how social distancing measures or kindergarten closures would be lifted.

A limitation of our study is its cross-sectional design. A longitudinal perspective is ideal to provide a better understanding of how the situation of families’ develops over time and whether concerns vary according to current restriction measures and number of COVID-19 cases. This longitudinal approach was provided in an analysis from Croatia, where surveys were performed in almost 1000 individuals on the day after the first patient was diagnosed with COVID-19 in the country and 3 weeks later.^12^ There, concerns and worries increased over the three weeks period as did the acceptance for the measures with increasing severity of the pandemic. Those who were at highest risk but also mothers represented the most concerned group^12^, consistent with earlier pandemics such as the swine flu.^13^ Therefore, parents, especially mothers, were the group we addressed in our study. While we did not investigate concerns longitudinally, Leipzig would rather compare to an early stage of the pandemic and Regensburg represents a situation where the pandemic has already increased its punch. Even though measures were almost identical in our two study locations, the course of the pandemic was different with much less and less severely affected individuals in Leipzig. Therefore, our study very nicely confirms and complements the findings from Croatia: Those results underline, when pandemic effects are at a low level, concerns may be less prominent and trust in the necessity of protective measures may decrease. To downplay concerns about the pandemic in a situation where one’s own experience with the disease and its impact are still limited, may even be a reasonable coping strategy. On the other hand, adherence to preventive measures over months is required to reinforce prevention for the duration of the pandemic.

We found differences in concerns and trust between our two study sites. However, we cannot differentiate between effects caused by differences in the course of the pandemic or the associated policy measures or by differences of the two regions. Also, residual confounding cannot be excluded.

Interestingly, it was especially found in Leipzig, that parents could also find some benefits in the current situation of lockdown. Significantly more often than in Regensburg, parents from Leipzig reported that they enjoyed spending more time together. Nevertheless, it must not be neglected that in some families, the risk for violence and vulnerability may increase for children during periods of school closures due to health emergencies.^14^

Our study population consisted mostly of well educated parents as we drew them from our ongoing pediatric cohort studies. This is a common observation as families with low education, low income and migration background are often underrepresented in longitudinal health studies.^15,16^ Due to this selection bias we may underestimate the impact of the pandemic on families who are underprivileged. However, we found similar tendencies in the two study populations and thus, the comparisons between the two regions are unlikely to be biased by these conditions.

Doubtless, the current COVID-19 pandemic is a stressor for many people across the world. Parents seem to be especially concerned and children are particularly vulnerable as they are in a critical phase of development. Spillover effects of parental stress towards children are well described, also for times of health crisis.^17^ What impact the current situation may have on mental health of the young generation is still to be determined but first studies suggest that indeed, they may be troublesome.^18^ Therefore, special concerns of families need to be addressed and support needs to be provided despite social distancing and measures of infection control.

In summary, most families with young children in our study were concerned about the COVID-19 pandemic and the impact of the associated lock-down measures, with major concerns directed towards economic situation and the lack of social contacts, but less towards their own health. Overall concerns, need for information on the pandemic and approval for federal measures were more pronounced in the more affected region. These data may suggest that more information about the extent and consequences of the pandemic, probably also on a more personal and individual scale, is needed in less affected regions to reassure people about the necessity and scale of preventive measures.

## Data Availability

Data Availability Statement: Data cannot be shared publicly because there exist ethical restrictions. The LIFE Child and KUNO Kids are a study collecting potentially sensitive information.
Publishing data sets is not covered by the informed consent provided by the study participants. Furthermore, the data protection concept of LIFE requests that all (external as well as internal) researchers interested in accessing data sign a project agreement. Researchers that are interested in accessing and analyzing data collected in the LIFE Child study may contact the data use and access committee. Researchers that are interested in accessing and analyzing data collected in the KUNO Kids study may contact the first author.

## Contributors

SB, TP, MV, MK and AK designed the study. SB, MV, CM, BSG, MM contributed to data collection. TP, MV and CM analysed the data. SB, TP, MV, CM, CA, WK, MK and AK contributed to data interpretation. SB, TP, MK and AK drafted the first version of the manuscript. All authors contributed to interpretation of data and critical revision of the manuscript.

## Ethics Approval

The LIFE Child study was approved by the Ethics Committe of the Medical Faculty of the University of Leipzig (Reg. No. 264/10-ek). The KUNO-Kids study was approved by the Ethics Committee of the University of Regensburg (Reference Number 14-101-0347).

## Data sharing

De-identified participant data of the KUNO Kids study will be made available upon reasonable request from MK. De-identified participant data of the LIFE Child study will be made available upon reasonable request from the LIFE Child data use and access committee.

## Acknowledgement

We thank all families for their participation.

The members of the KUNO Kids study group are: Andreas Ambrosch (Institute of Laboratory Medicine, Microbiology and Hygiene, Barmherzige Brüder Hospital, Regensburg, Germany), Petra Arndt (ZNL Transfercenter of Neuroscience and Learning, University of Ulm, Ulm, Germany), Andrea Baessler (Department of Internal Medicine II, Regensburg University Medical Center, Regensburg, Germany), Mark Berneburg (Department of Dermatology, University Medical Centre Regensburg, Regensburg, Germany), Stephan Böse-O’Reilly (University Children’s Hospital Regensburg (KUNO), Hospital St. Hedwig of the Order of St. John, Regensburg, Germany), Romuald Brunner (Clinic of Child and Adolescent Psychiatry, Psychosomatics and Psychotherapy, Bezirksklinikum Regensburg (medbo), Regensburg, Germany), Wolfgang Buchalla (Department of Conservative Dentistry and Periodontology, University Hospital Regensburg, University of Regensburg, Regensburg, Germany), Sara Fill Malfertheiner (Clinic of Obstetrics and Gynecology St. Hedwig, University of Regensburg, Regensburg, Germany), André Franke (Institute of Clinical Molecular Biology, Christian-Albrechts-University of Kiel, Kiel, Germany), Sebastian Häusler (Clinic of Obstetrics and Gynecology St. Hedwig, University of Regensburg, Regensburg, Germany), Iris Heid (Department of Genetic Epidemiology, University of Regensburg, Regensburg, Germany), Caroline Herr (Bavarian Health and Food Safety Authority (LGL), Munich, Germany), Wolfgang Högler (Department of Pediatrics and Adolescent Medicine, Johannes Kepler University Linz, Linz, Austria), Sebastian Kerzel (Department of Pediatric Pneumology and Allergy, University Children’s Hospital Regensburg (KUNO), Hospital St. Hedwig of the Order of St. John, Regensburg, Germany), Michael Koller (Center for Clinical Studies, University Hospital Regensburg, Regensburg, Germany), Michael Leitzmann (Department of Epidemiology and Preventive Medicine, University of Regensburg, Regensburg, Germany), David Rothfuß (City of Regensburg, Coordinating Center for Early Interventions, Regensburg, Germany), Wolfgang Rösch (Department of Pediatric Urology, University Medical Center, Regensburg, Germany), Bianca Schaub (Pediatric Allergology, Department of Pediatrics, Dr. von Hauner Children’s Hospital, University Hospital, LMU Munich, Munich, Germany), Bernhard H.F. Weber (Institute of Human Genetics, University of Regensburg, Regensburg, Germany), Stephan Weidinger (Department of Dermatology, Venereology and Allergy, University Hospital Schleswig-Holstein, Campus Kiel, Kiel, Germany) and Sven Wellmann (Department of Neonatology, University Children’s Hospital Regensburg (KUNO), Hospital St. Hedwig of the Order of St. John, Regensburg, Germany).

## What is already known on this topic?

- At the beginning of the COVID-19 pandemic, children were assumed to be not seriously affected by SARS-CoV-2.
- Then it was realized, that the pandemic and the associated lockdown-measures might have impact on the psychosocial situation of families and children.
- Studies investigating samples of children/families from the general population using primary data were lacking.

## What this study adds?

- Investigating two population based childhood cohorts in Germany we found that many families were concerned about the COVID-19 pandemic and associated lockdown measures
- Families form regions with a lower prevalence of COVID-19 showed less concerns but also less trust in political measures.

